# Burden and trend of kidney cancer attributable to high body mass index among middle-aged males in East and Southeast Asia from 1990 to 2021

**DOI:** 10.1101/2025.08.26.25334520

**Authors:** Zhengkun Wang, Yikai Wang, Qiwei Wang, Yi Rong, Weibing Shuang

## Abstract

**Background:** With economic development and lifestyle transitions, obesity prevalence has increased, and kidney cancer is significantly influenced by high body mass index (BMI), particularly among middle-aged males. This study analyzes the burden and trends of kidney cancer attributable to high BMI among middle-aged males in East and Southeast Asia, and projects future trends.

**Methods:** Based on the Global Burden of Disease (GBD) 2021 study, we assessed the burden of kidney cancer attributable to high BMI among middle-aged males in East and Southeast Asia from 1990 to 2021, specifically comparing deaths, disability-adjusted life years (DALYs), age-standardized DALY rates (ASDR), and age-standardized mortality rates (ASMR). Furthermore, we analyzed spatiotemporal trends of kidney cancer at multiple levels. Additionally, the Bayesian Age-Period-Cohort (BAPC) model was employed to predict future burden trends through 2040.

**Results:** From 1990 to 2021, DALYs and deaths attributable to high BMI-induced kidney cancer among middle-aged males in East Asia increased dramatically by 530.73% and 517.28%, respectively, while corresponding increases in Southeast Asia reached 478.68% and 478.70%. These growth rates significantly exceeded the global average. In 2021, the ASDR in East Asia was 15.63 per 100,000, and the ASMR was 0.40 per 100,000. For Southeast Asia, the ASDR and ASMR were 9.40 per 100,000 and 0.25 per 100,000, respectively, highlighting substantial regional disparities. Both regions exhibited a positive correlation between Socio-demographic Index (SDI) and ASDR/ASMR, with a disproportionately higher burden observed in the 55–59-year age group. Projections indicate that the burden in these regions will continue to rise through 2040.

**Conclusion:** The increasing burden of kidney cancer attributable to high BMI among middle-aged males in East and Southeast Asia has become a serious public health challenge. Developing and implementing targeted interventions to address the escalating obesity epidemic and its impact on kidney cancer have become critically urgent.

## Introduction

Kidney cancer represents a significant global health challenge, with a notably higher burden among men, ranking as the ninth most common malignancy in males. Due to its poor prognosis, the 5-year survival rate for advanced metastatic cases is only 12% ^[1–3]^. Key risk factors include high body mass index (BMI), smoking, and occupational exposure to trichloroethylene. Notably, high BMI is increasingly recognized as a critical risk factor for tumor development ^[4]^. According to the 2019 Global Burden of Disease (GBD) Study, high BMI accounted for 19.05% of global kidney cancer deaths, surpassing the contribution of smoking (18.09%) ^[5]^. Epidemiological studies consistently demonstrate a positive correlation between BMI and kidney cancer incidence, with overweight and obesity increasing risk by 35% and 76%, respectively. This association may be attributed to dysregulated adipokine secretion, insulin resistance, and altered sex hormone activity ^[6]^.

In 2022, approximately 890 million people globally were living with obesity (representing 16% of the world’s population), with an adult obesity prevalence of 43% ^[7]^. Asia, a continent marked by significant heterogeneity in lifestyles, economic conditions, and disease epidemiology, has experienced a rising trend in high BMI prevalence among males, particularly in East and Southeast Asia. This surge is closely linked to regional economic development, urbanization, and lifestyle shifts—characterized by the adoption of Western diets and sedentary behaviors ^[8–10]^. Notably, middle-aged males face distinct challenges, including work-related stress, social engagements involving high-fat diets, reduced physical activity, and declining sex hormone levels, which collectively drive elevated obesity rates in this demographic ^[11]^.

However, while prior research has predominantly focused on the burden of kidney cancer in older adults with high BMI ^[12]^, a critical gap exists regarding middle-aged males in Asia. This demographic exhibits rising obesity rates linked to work stress, social engagements, high-fat diets, reduced physical activity, and declining sex hormones ^[11]^. The GBD study provides the most comprehensive epidemiological assessment of kidney cancer to date ^[13]^. Therefore, utilizing the latest GBD 2021 data, this study systematically analyzes the kidney cancer burden attributable to high BMI among middle-aged males (45–59 years, WHO classification) in East and Southeast Asia, aiming to inform targeted prevention strategies in this region.

## Materials and methods

### Data sources

Data for this study were obtained from the GBD 2021 database (https://vizhub.healthdata.org/gbd-results/, accessed on June 23, 2025). The study analyzed indicators associated with KC, including Disability-Adjusted Life Years (DALYs) and Deaths, across East and Southeast Asia from 1990 to 2021. The data were stratified by location, Socio-demographic Index (SDI), and age group (45–49, 50–54, 55–59 years) to examine spatiotemporal patterns in the disease burden among middle-aged males with high BMI. DALYs are used as a metric to measure the total health loss caused by specific diseases or risk factors in population health. The SDI quantifies the socio-economic status of a country or location, reflecting its association with health outcomes. The SDI scale ranges from 0 (indicating high fertility, low education, and low income) to 1 (low fertility, high education, and high income) ^[14]^. Since the GBD database is publicly accessible and contains no personally identifiable or sensitive data, and its source studies were approved by their respective institutional review boards, this study was exempt from ethics review.

### Statistical analysis

Kidney cancer primarily affects patient quality of life and is associated with direct mortality. The crude rates (DALYs and deaths), obtained in this study, provide fundamental metrics to measure epidemiological trends; however, differences in population age distribution may bias burden estimates. To enhance comparability, crude rates were adjusted by applying age-specific weights based on the age composition of a standard population, resulting in the age-standardized rate (ASR). The age-standardized DALY rate (ASDR) and age-standardized mortality rate (ASMR) were used to calculate the Estimated Annual Percentage Change (EAPC), reflecting annual trends of kidney cancer burden from 1990 to 2021. Rates are expressed as age-standardized values per 100,000 population, with 95% uncertainty intervals (UIs).

In this study, temporal trends of ASDR and ASMR for kidney cancer from 1990 to 2021 were analyzed using the Joinpoint software (version 5.1.0). The annual percent change (APC) and average annual percentage change (AAPC) were calculated to quantify trend direction and statistical significance. Trends were classified as “decreasing” or “increasing” when the APC and AAPC were statistically distinct from zero ^[15]^.

We employed the Bayesian age-period-cohort (BAPC) model to project the future burden of kidney cancer among middle-aged males with high BMI. This model, accessible via the BAPC R package, integrates age, period, and cohort effects within a comprehensive Bayesian framework. For Bayesian inference, we utilized the Integrated Nested Laplace Approximation (INLA) method. INLA efficiently approximates the marginal posterior distributions of model parameters, providing a computationally efficient alternative to conventional Markov Chain Monte Carlo (MCMC) methods ^[16]^.

By analyzing data from 1990 to 2021, this study quantifies temporal trends in each epidemiologic metric and systematically characterizes, through descriptive statistics and data visualization techniques, the evolution of kidney cancer epidemiology among middle-aged males across geographic, age-specific, and temporal dimensions. All analyses were conducted using R version 4.3.3.

## Results

### The burden of kidney cancer attributable to high BMI among middle-aged males in East and Southeast Asia

From 1990 to 2021, the burden of kidney cancer attributable to high BMI among middle-aged males increased substantially in both East and Southeast Asia. In East Asia, the number of DALYs surged from 4,488.76 (95% UI: 1,726.19–7,730.99) to 28,312.10 (95% UI: 10,802.67–50,778.11), representing a striking 530.73% increase. Similarly, deaths rose from 118.40 (95% UI: 45.59–203.83) to 730.86 (95% UI: 277.43–1,316.02), marking a 517.28% increase. Conversely, in Southeast Asia, the DALYs increased from 953.49 (95% UI: 352.34–1,624.58) to 5,517.63 (95% UI: 2,063.33–9,069.72), a 478.68% rise. Likewise, deaths climbed from 24.93 (95% UI: 9.18–42.54) to 144.27 (95% UI: 53.74–237.00), reflecting a 478.70% increase. Notably, the ASR demonstrated divergent trends. In East Asia, the ASDR per 100,000 population escalated from 5.72 (95% UI: 2.20–9.85) to 15.63 (95% UI: 5.96–28.01), with an EAPC of 3.772 (95% CI: 3.538–4.006). Correspondingly, the ASMR increased from 0.15 (95% UI: 0.06–0.26) to 0.40 (95% UI: 0.15–0.72), at an EAPC of 3.667 (95% CI: 3.442–3.892). By contrast, Southeast Asia exhibited lower growth in ASR: The ASDR rose from 4.22 (95% UI: 1.56–7.19) to 9.40 (95% UI: 3.52–15.45), with an EAPC of 2.317 (95% CI: 2.115–2.519), while the ASMR increased from 0.11 (95% UI: 0.04–0.19) to 0.25 (95% UI: 0.09–0.40), at an EAPC of 2.314 (95% CI: 2.113–2.516) (Table 1 and Table 2).

**Table 1.** Age-standardized DALYs rates and corresponding EAPC of kidney cancer attributable to high BMI among middle-aged males in East and Southeast Asia in 1990 and 2021.

| Location | 1990 |  | 2021 |  | 1990-2021 |
| --- | --- | --- | --- | --- | --- |
|  | DALYs | ASDR | DALYs | ASDR | EAPC |
|  | n (95% UI) | per 100,000<br>n (95% UI) | n (95% UI) | per 100,000<br>n (95% UI) | n (95% CI) |
| Global | 70501.32<br>(27471.38-114303.89) | 22.12<br>(8.62-35.86) | 164242.21<br>(67185.75-268411.90) | 24.95<br>(10.2-40.76) | 0.377<br>(0.189,0.566) |
| East Asia | 4488.76 (1726.19-7730.99) | 5.72 (2.20-9.85) | 28312.10 (10802.67-50778.11) | 15.63 (5.96-28.01) | 3.772 (3.538,4.006) |
| China | 4330.36 (1657.53-7504.72) | 5.72 (2.19-9.92) | 27276.85 (10351.16-49266.50) | 15.54 (5.89-28.05) | 3.778 (3.534,4.023) |
| Taiwan (Province of China) | 106.19 (39.15-182.06) | 8.16 (3.01-13.99) | 871.39 (326.03-1531.76) | 31.82 (11.92-55.86) | 3.933 (3.331,4.537) |
| North Korea | 52.21 (16.90-109.40) | 3.47 (1.13-7.27) | 163.86 (55.00-336.02) | 5.72 (1.91-11.72) | 1.638 (1.575,1.701) |
| South Korea | 263.54 (98.93-474.13) | 9.25 (3.47-16.67) | 922.74 (345.46-1657.20) | 13.71 (5.13-24.61) | 1.135 (0.669,1.604) |
| Japan | 1619.80 (610.54-2666.34) | 12.85 (4.85-21.16) | 1809.10 (697.84-2913.54) | 13.23 (5.10-21.31) | 0.342 (0.043,0.641) |
| Southeast Asia | 953.49 (352.34-1624.58) | 4.22 (1.56-7.19) | 5517.63 (2063.33-9069.72) | 9.40 (3.52-15.45) | 2.317 (2.115,2.519) |
| Sri Lanka | 165.10 (51.03-331.17) | 17.20 (5.29-34.52) | 152.40 (47.20-322.59) | 7.98 (2.47-16.89) | -4.873 (-6.027,-3.705) |
| Timor-Leste | 0.39 (0.12-0.81) | 1.16 (0.36-2.44) | 2.37 (0.65-5.19) | 3.36 (0.92-7.32) | 3.903 (3.653,4.154) |
| Cambodia | 10.61 (3.56-22.08) | 2.94 (0.99-6.12) | 72.52 (22.74-149.46) | 6.83 (2.14-14.07) | 2.959 (2.845,3.074) |
| Thailand | 169.36 (51.35-343.23) | 5.15 (1.56-10.44) | 1252.26 (445.72-2388.65) | 15.82 (5.64-30.21) | 3.738 (3.601,3.874) |
| Laos | 5.63 (1.81-11.95) | 3.23 (1.04-6.85) | 35.43 (10.35-74.84) | 8.00 (2.34-16.88) | 3.205 (3.061,3.349) |
| Malaysia | 73.68 (25.60-138.93) | 8.89 (3.10-16.73) | 334.94 (112.03-648.60) | 14.09 (4.71-27.28) | 1.223 (1.037,1.409) |
| Mauritius | 4.50 (1.68-7.77) | 7.95 (2.97-13.74) | 23.45 (9.23-40.23) | 17.24 (6.76-29.54) | 0.049 (-1.514,1.638) |
| Brunei Darussalam | 1.67 (0.55-3.13) | 15.80 (5.22-29.66) | 12.72 (4.33-24.09) | 33.72 (11.48-63.92) | 3.163 (2.959,3.368) |
| Indonesia | 260.81 (93.70-483.41) | 2.78 (1.00-5.15) | 2108.42 (754.28-3835.16) | 8.72 (3.12-15.86) | 3.910 (3.759,4.062) |
| Maldives | 0.37 (0.12-0.85) | 3.52 (1.09-8.05) | 2.65 (0.93-5.32) | 6.22 (2.21-12.41) | 1.687 (1.570,1.805) |
| Singapore | 23.33 (8.42-40.55) | 12.33 (4.45-21.40) | 90.72 (34.88-157.52) | 13.37 (5.14-23.21) | 0.330 (-0.143,0.805) |
| Philippines | 184.09 (65.18-316.07) | 6.62 (2.34-11.36) | 1032.89 (365.91-1837.8) | 13.45 (4.76-23.92) | 2.202 (2.050,2.355) |
| Myanmar | 56.46 (18.41-123.59) | 2.92 (0.95-6.39) | 227.10 (74.28-442.30) | 5.63 (1.84-10.96) | 2.178 (2.126,2.229) |
| Seychelles | 0.52 (0.18-0.98) | 14.10 (4.93-26.37) | 2.74 (0.93-5.10) | 24.46 (8.32-45.58) | 0.863 (0.136,1.595) |
| Viet Nam | 20.58 (6.97-40.62) | 0.74 (0.25-1.46) | 262.77 (80.54-549.78) | 3.02 (0.93-6.31) | 5.531 (5.201,5.863) |

DALYs, disability-adjusted life years; ASDR, age-standardized DALY rates; ASMR,
age-standardized mortality rates; EAPC, estimated annual percentage change.

**Table 2.** Age-standardized Deaths rates and corresponding EAPC of kidney cancer attributable to high BMI among middle-aged males in East and Southeast Asia in 1990 and 2021.

**Asia in 1990 and 2021**
| Location | 1990 |  | 2021 |  | 1990-2021 |
| --- | --- | --- | --- | --- | --- |
|  | Deaths | ASMR | Deaths | ASMR | EAPC |
|  | n (95% UI) | per 100,000<br>n (95% UI) | n (95% UI) | per 100,000<br>n (95% UI) | n (95% CI) |
| Global | 1863.71<br>(724.68-3027.13) | 0.58<br>(0.23-0.95) | 4293.54<br>(1757.96-7041.71) | 0.65<br>(0.27-1.07) | 0.349<br>(0.167,0.530) |
| East Asia | 118.40 (45.59-203.83) | 0.15 (0.06-0.26) | 730.86 (277.43-1316.02) | 0.40 (0.15-0.72) | 3.667 (3.442,3.892) |
| China | 114.30 (43.79-197.92) | 0.15 (0.06-0.26) | 704.26 (265.87-1277.14) | 0.40 (0.15-0.72) | 3.670 (3.435,3.906) |
| Taiwan (Province of China) | 2.75 (1.01-4.69) | 0.21 (0.08-0.36) | 22.35 (8.25-39.58) | 0.81 (0.30-1.43) | 3.855 (3.256,4.457) |
| North Korea | 1.35 (0.44-2.84) | 0.09 (0.03-0.19) | 4.26 (1.44-8.74) | 0.15 (0.05-0.30) | 1.618 (1.560,1.675) |
| South Korea | 6.95 (2.59-12.53) | 0.25 (0.09-0.44) | 24.10 (9.01-43.05) | 0.36 (0.13-0.63) | 1.046 (0.586,1.507) |
| Japan | 43.25 (16.23-71.29) | 0.34 (0.13-0.56) | 46.89 (18.06-75.53) | 0.34 (0.13-0.55) | 0.276 (-0.017,0.571) |
| Southeast Asia | 24.93 (9.18-42.54) | 0.11 (0.04-0.19) | 144.27 (53.74-237.00) | 0.25 (0.09-0.40) | 2.314 (2.113,2.516) |
| Sri Lanka | 4.31 (1.32-8.65) | 0.45 (0.14-0.91) | 3.97 (1.22-8.37) | 0.21 (0.06-0.44) | -4.910 (-6.065,-3.740) |
| Timor-Leste | 0.01 (0.00-0.02) | 0.03 (0.01-0.06) | 0.06 (0.02-0.13) | 0.09 (0.02-0.19) | 3.885 (3.637,4.133) |
| Cambodia | 0.28 (0.09-0.59) | 0.08 (0.03-0.16) | 1.92 (0.60-3.96) | 0.18 (0.06-0.37) | 2.929 (2.816,3.042) |
| Thailand | 4.42 (1.34-8.92) | 0.13 (0.04-0.27) | 32.46 (11.49-61.78) | 0.41 (0.14-0.78) | 3.687 (3.555,3.820) |
| Laos | 0.15 (0.05-0.32) | 0.09 (0.03-0.18) | 0.94 (0.28-1.98) | 0.21 (0.06-0.45) | 3.198 (3.052,3.345) |
| Malaysia | 1.94 (0.68-3.64) | 0.24 (0.08-0.44) | 8.81 (2.93-16.96) | 0.37 (0.12-0.71) | 1.188 (1.001,1.376) |
| Mauritius | 0.12 (0.04-0.20) | 0.21 (0.08-0.36) | 0.62 (0.24-1.06) | 0.45 (0.18-0.77) | -0.001 (-1.560,1.582) |
| Brunei Darussalam | 0.04 (0.01-0.08) | 0.42 (0.14-0.79) | 0.33 (0.11-0.63) | 0.89 (0.30-1.68) | 3.107 (2.903,3.312) |
| Indonesia | 6.82 (2.46-12.62) | 0.07 (0.03-0.14) | 55.19 (19.80-100.63) | 0.23 (0.08-0.42) | 3.919 (3.768,4.070) |
| Maldives | 0.01 (0.00-0.02) | 0.09 (0.03-0.22) | 0.07 (0.02-0.14) | 0.16 (0.06-0.32) | 1.557 (1.436,1.678) |
| Singapore | 0.62 (0.22-1.08) | 0.33 (0.12-0.57) | 2.38 (0.91-4.10) | 0.35 (0.13-0.60) | 0.228 (-0.239,0.697) |
| Philippines | 4.77 (1.69-8.20) | 0.17 (0.06-0.3) | 27.03 (9.53-48.06) | 0.35 (0.12-0.63) | 2.233 (2.078,2.388) |
| Myanmar | 1.50 (0.49-3.29) | 0.08 (0.03-0.17) | 6.02 (1.97-11.72) | 0.15 (0.05-0.29) | 2.175 (2.127,2.223) |
| Seychelles | 0.01 (0.00-0.03) | 0.37 (0.13-0.70) | 0.07 (0.02-0.13) | 0.65 (0.22-1.20) | 0.829 (0.104,1.559) |
| Viet Nam | 0.55 (0.19-1.09) | 0.02 (0.01-0.04) | 6.90 (2.13-14.48) | 0.08 (0.02-0.17) | 5.485 (5.156,5.815) |

DALYs, disability-adjusted life years; ASDR, age-standardized DALY rates; ASMR, age-standardized mortality rates; EAPC, estimated annual percentage change.

### The burden of kidney cancer attributable to high BMI among middle-aged males in East and Southeast Asia

At the national level across East and Southeast Asia in 2021, China had the highest burden of kidney cancer DALYs and deaths attributable to high BMI among middle-aged males, followed by Japan. Notably, Brunei Darussalam, Taiwan (Province of China), and Seychelles showed the highest ASDR and ASMR. Meanwhile, Viet Nam exhibited the most rapid increases in ASDR and ASMR, with respective EAPCs of 5.531 (95% CI: 5.201–5.863) and 5.485 (95% CI: 5.156–5.815).

In contrast, Mauritius remained relatively stable, with EAPCs of 0.049 (95% CI: -1.514–1.638) for ASDR and -0.001 (95% CI: -1.560–1.582) for ASMR. However, Sri Lanka was the only country demonstrating significant declines, with EAPCs of -4.873 (95% CI: -6.027 to -3.705) for ASDR and -4.910 (95% CI: -6.065 to -3.740) for ASMR. It is noteworthy that during 2002–2005, Mauritius, Sri Lanka, and Seychelles experienced substantial reductions in both ASDR and ASMR (Fig1, Fig2 and S1 Table).

**Fig 1.**
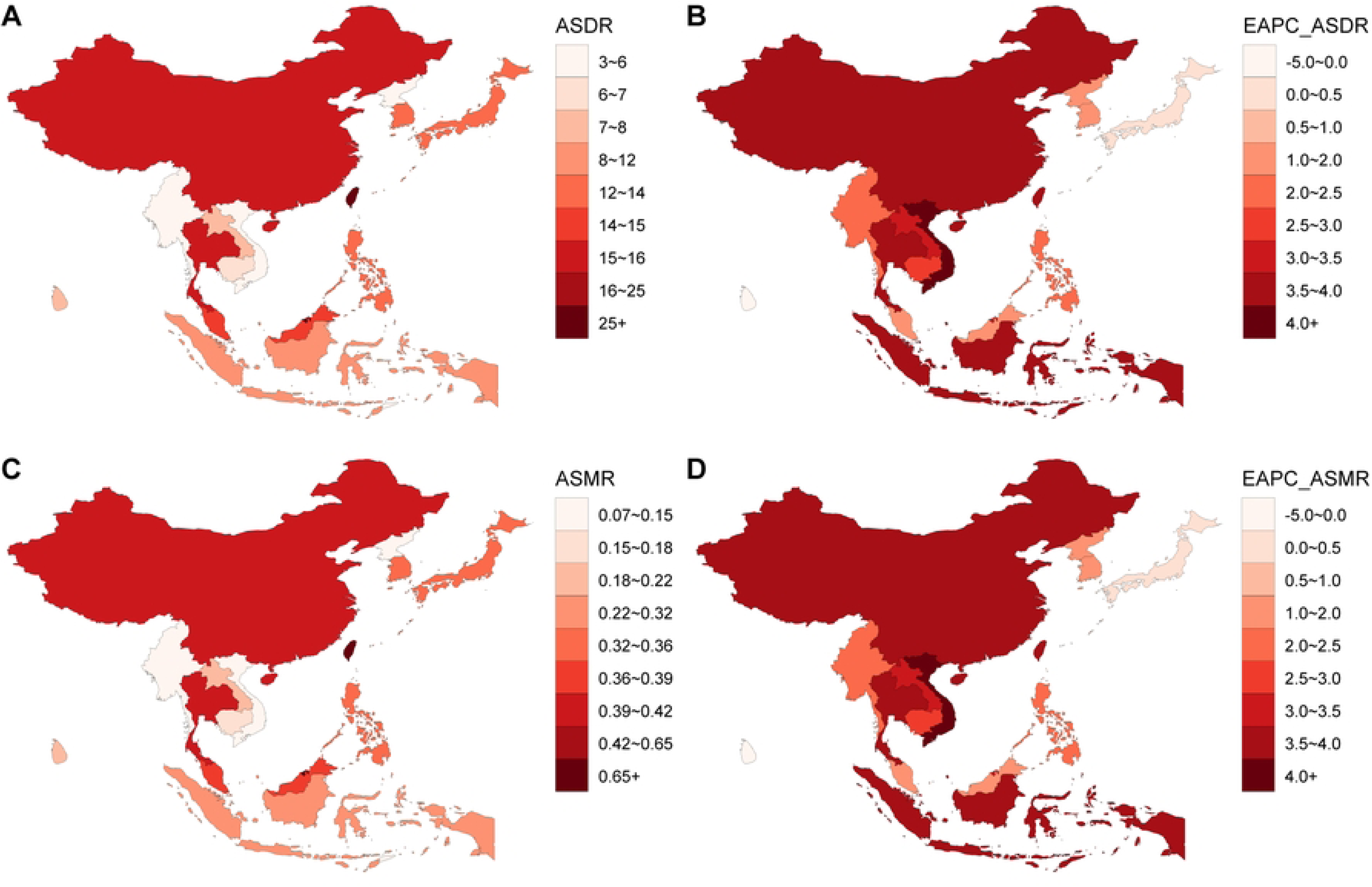
The burden of kidney cancer attributable to high BMI among middle-aged males in East and Southeast Asia. (A) The ASDR in 2021; (B) The ASMR in 2021; (C) The EAPC of ASDR, 1990–2021; (D) The EAPC of ASMR, 1990–2021. DALYs, disability-adjusted life years; ASDR, age-standardized DALY rates; ASMR, age-standardized mortality rates; EAPC, estimated annual percentage change.

**Fig 2.**
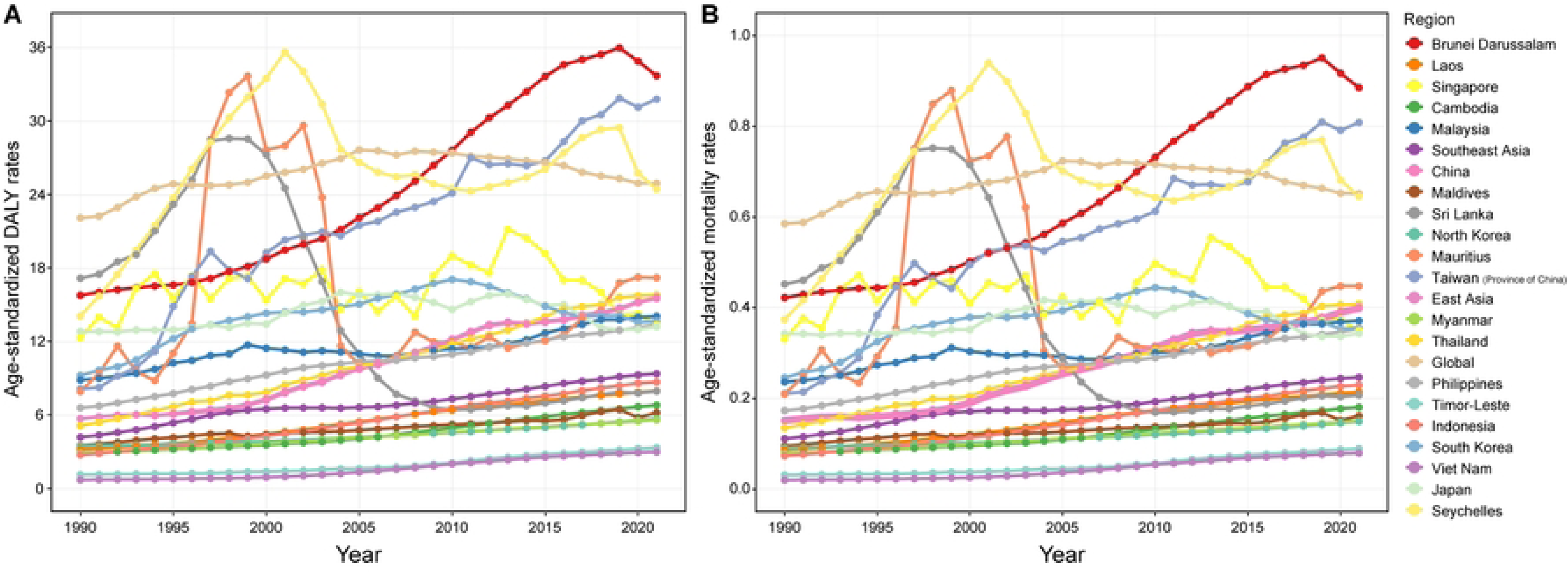
The burden of of kidney cancer attributable to high BMI among middle-aged males in East and Southeast Asia. (A) ASDR, 1990–2021; (B) ASMR, 1990–2021. ASDR, age-standardized DALY rates; ASMR, age-standardized mortality rates.

### Socio-demographic disparities in the burden of kidney cancer attributable to high BMI among middle-aged males in East and Southeast Asia

At the national level in East and Southeast Asia from 1990 to 2021, a strong positive correlation was observed between the SDI and the ASDR of kidney cancer burden among middle-aged males (Spearman’s ρ = 0.79, P < 0.001), indicating that ASDR increased with rising SDI. Similarly, the ASMR exhibited an identical correlation pattern (ρ = 0.79, P < 0.001). Furthermore, this positive association persisted in 2021 between national SDI and ASDR (ρ = 0.71, P < 0.001), with ASMR showing a comparable trend (ρ = 0.71, P < 0.001). Geographically, regions including Brunei Darussalam, Taiwan (Province of China), Seychelles, Thailand, Philippines, and Laos demonstrated higher-than-expected ASDR in 2021, whereas countries such as Malaysia, South Korea, Singapore, Japan, Sri Lanka, and Vietnam had lower-than-expected ASDR (Fig 3).

**Fig 3.**
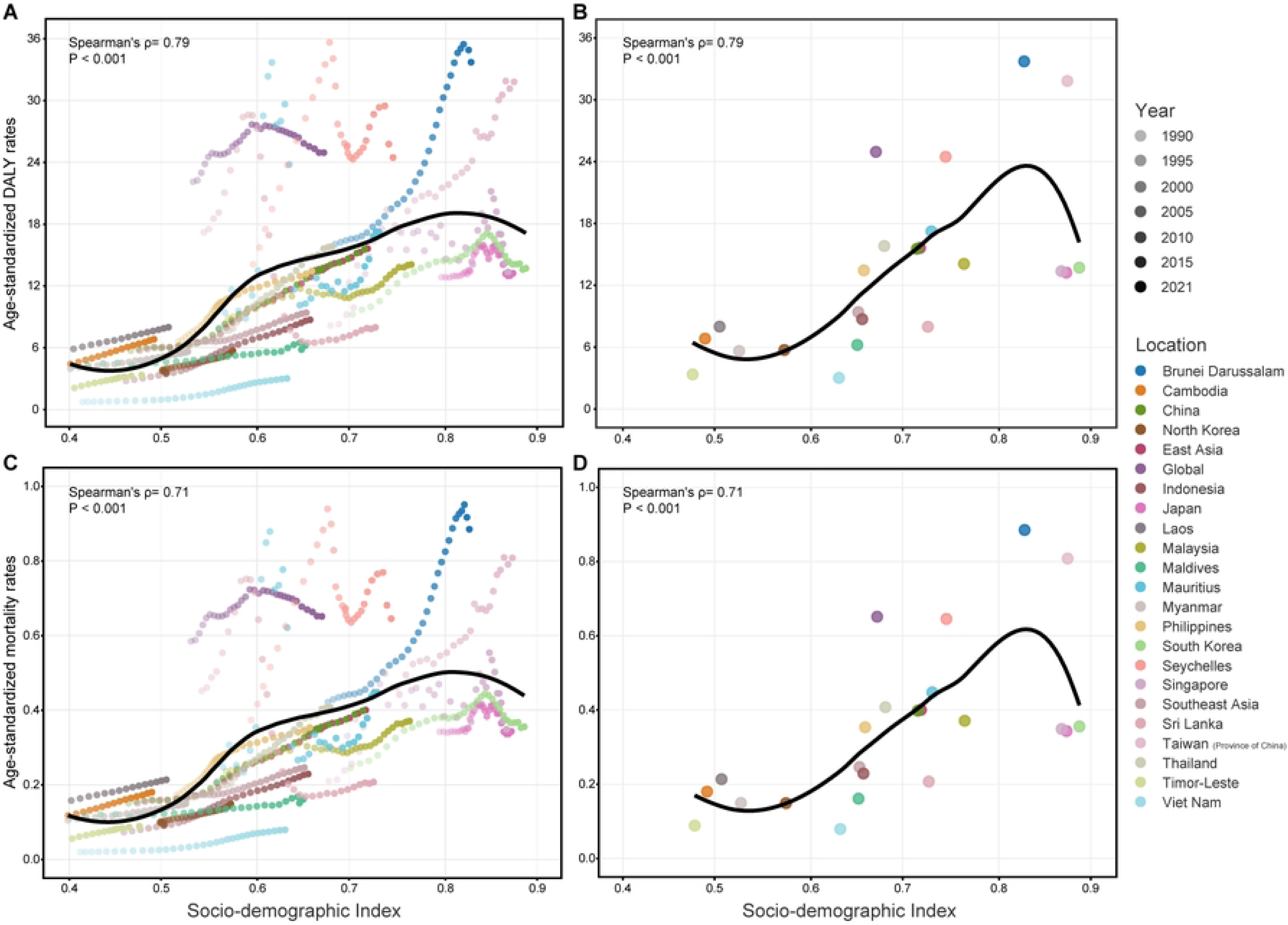
Socio-demographic disparities in the burden of kidney cancer attributable to high BMI among middle-aged males in East and Southeast Asia. (A) ASDR by SDI, 1990–2021; (B) ASDR by SDI in 2021; (C) ASMR by SDI, 1990–2021; (D) ASMR by SDI in 2021. ASDR, age-standardized DALY rate; ASMR, age-standardized mortality rates; SDI, sociodemographic index.

### Age-specific trends in the burden of kidney cancer attributable to high BMI among middle-aged males in East and Southeast Asia

A comprehensive joinpoint analysis revealed distinct temporal trends in DALY and mortality rates for kidney cancer attributable to high BMI among males aged 45–59 years in East and Southeast Asia from 1990 to 2021. Notably, both regions exhibited estimated AAPC for DALY and mortality rates exceeding global averages, with disease burden escalating with age. East Asia consistently demonstrated higher baseline burdens and AAPC values than Southeast Asia across all age groups.

In East Asia, DALY rate AAPC values declined with advancing age: 3.506 (45–49), 3.357 (50–54), and 3.025 (55–59), while mortality rates AAPC showed a parallel trend (3.430, 3.281, and 2.943, respectively). The 45–49 age group experienced sharp DALY rate increases during 1999–2004 and 2007–2012, followed by a slow decline from 2012 to 2018 and a subsequent rebound. Conversely, the 50– 54 group exhibited a rapid increase from 1999 to 2012, interrupted by a brief decline during 2012–2015 before rebounding. In contrast, the 55–59 group maintained a steady upward trajectory that accelerated during 1996–2001 and stabilized after 2015 (Fig 4 and S2 Table).

**Fig 4.**
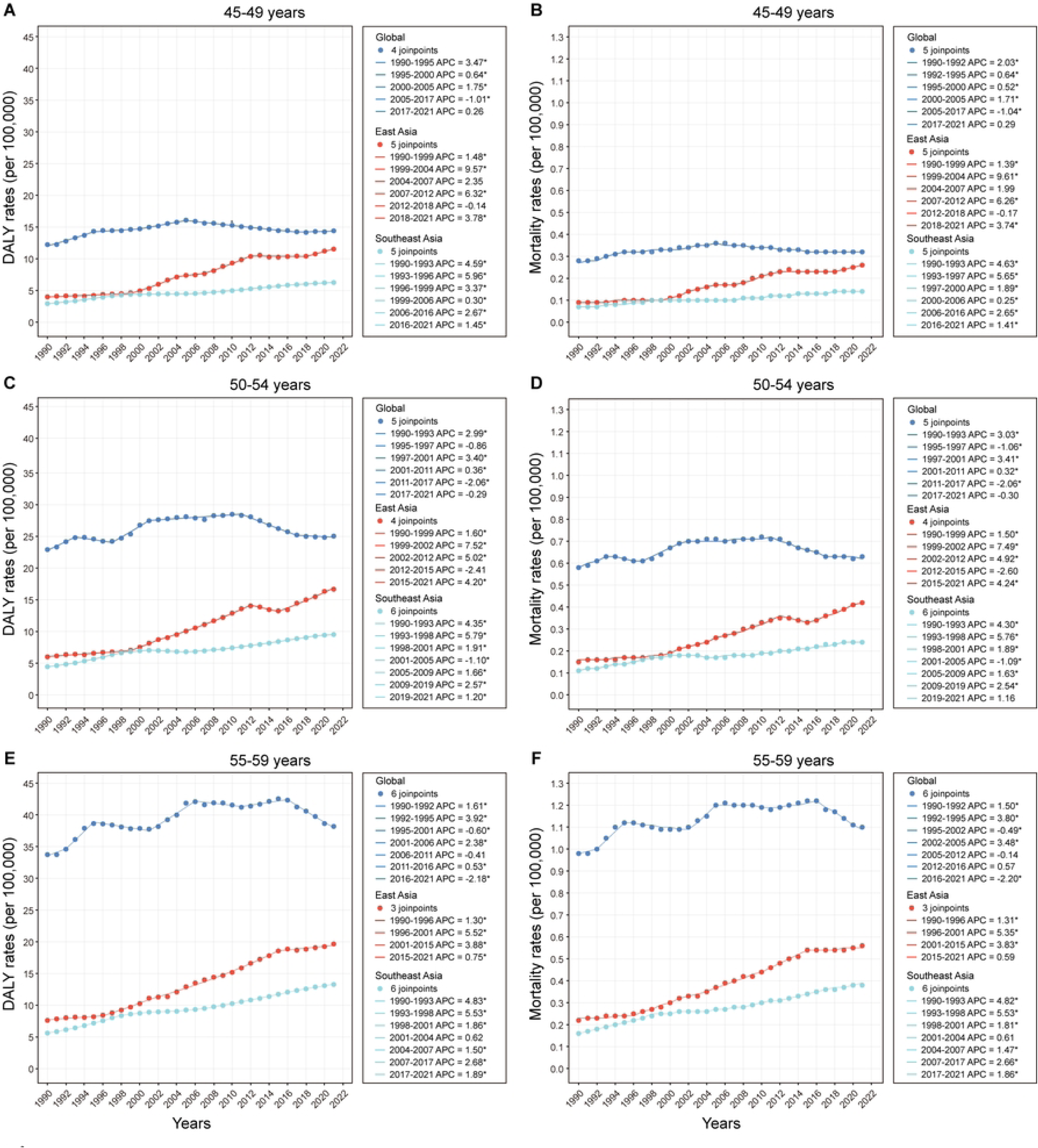
Age-specific trends in the burden of kidney cancer attributable to high BMI among middle-aged males. (A-B) 45–49 age group; (C-D) 50–54 age group; (E-F) 55–59 age group. APC: annual percent change.

Conversely, in Southeast Asia, DALY rate AAPC increased with advancing age: 2.496 (45–49), 2.497 (50–54), and 2.842 (55–59), with mortality rate AAPC mirroring this pattern (2.474, 2.479, and 2.822, respectively). The 45–49 age group exhibited steep increases during 1990–1997, remained stable from 2000 to 2006, and resumed growth thereafter. Similarly, the 50–54 group rose rapidly from 1990 to 1998, declined during 2001–2005, and stabilized subsequently. In contrast, the 55–59 group maintained a steadily upward trajectory overall, with accelerated growth during 1990–1998 and stabilization between 2001–2004 (Fig 4 and S2 Table).

### Projected trends in the burden of kidney cancer attributable to high BMI among middle-aged males in East and Southeast Asia

From 1990 to 2021, the burden of kidney cancer attributable to high BMI among middle-aged males in East and Southeast Asia, as measured by ASDR and ASMR, showed a significant increase. Projections indicate that both DALYs and deaths will continue to rise from 2022 to 2040. Specifically, in East Asia, DALYs are projected to reach 43,809.19 by 2040 (57.85% higher than 2021), with deaths rising to 948.77 (34.14% increase). In Southeast Asia, DALYs are expected to increase to 9,134.45 (64.55% higher) and deaths to 262.78 (80.24% higher), indicating more pronounced absolute growth. For age-standardized rates, ASDR in East Asia will rise to 25.53 per 100,000 (63.38% increase), while ASMR will reach 0.55 per 100,000 (38.84% increase). In Southeast Asia, ASDR will climb to 12.46 per 100,000 (32.21% increase) and ASMR to 0.36 per 100,000 (44.81% increase). Notably, although East Asia exhibits a higher ASDR growth rate, Southeast Asia shows a faster ASMR increase. Overall, both regions’ disease burden growth rates significantly exceed the global average (Fig 5 and S3 Table).

**Fig 5.**
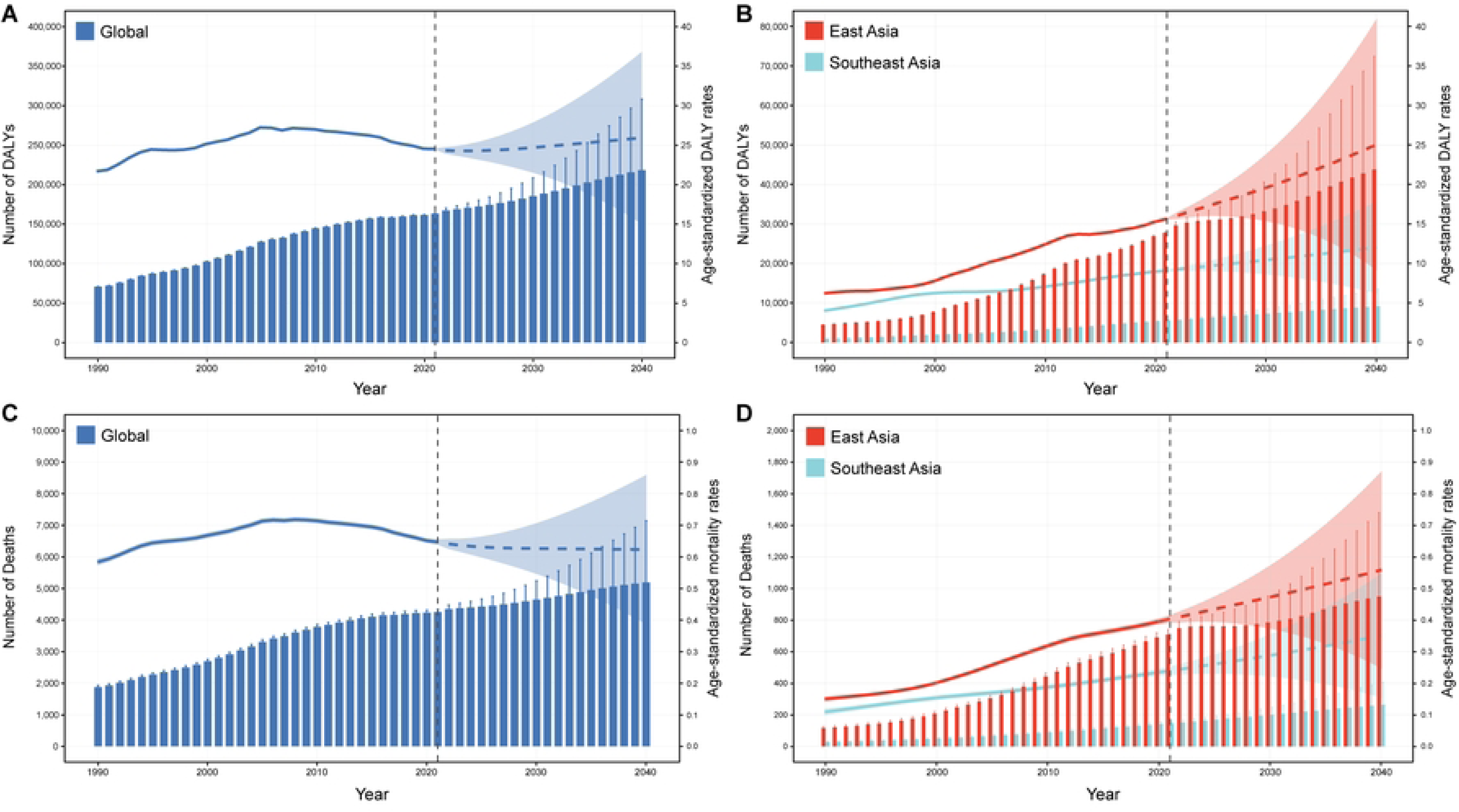
Projected trends in the burden of kidney cancer attributable to high BMI among middle-aged male, 2022–2040. (A) Global DALY numbers and ASDR; (B) DALY numbers and ASDR in East and Southeast Asia; (C) Global death numbers and ASMR; (D) Death numbers and ASMR in East and Southeast Asia. ASDR, age-standardized DALY rate; ASMR, age-standardized mortality rates.

## Discussion

Kidney cancer ranks as the ninth most common malignancy in males, with 25%–60% of cases being asymptomatic and detectable only incidentally through imaging ^[17]^. While surgery offers the best prognosis for early-stage disease, advanced kidney cancer carries a poor outlook, underscoring the critical importance of preventive strategies ^[18]^. Notably, obesity-induced dysfunctional adipose tissue drives alterations in serum adipokine levels, a key mechanism in obesity-related tumorigenesis ^[19]^. Obesity also promotes chronic low-grade systemic inflammation, characterized by activation of acute-phase response proteins and systemic inflammatory mediators ^[20]^. Furthermore, obesity is a major risk factor for insulin resistance, which triggers hyperinsulinemia and elevated insulin-like growth factor-1 (IGF-1)—both established drivers of cancer development ^[21]^. Additionally, oxidative stress, lipid peroxidation, renal hypoxia, tumor suppressor gene inactivation, and hypertension may collectively contribute to obesity-associated kidney cancer pathogenesis ^[22]^.

Notably, rapid global economic development and shifts in dietary habits have fueled the obesity epidemic, with approximately 2 billion people overweight worldwide ^[23]^, one-third of whom are classified as obese. Crucially, Asian populations exhibit 3%–5% higher body fat at equivalent BMI levels compared to Caucasians ^[24]^. Utilizing the latest GBD 2021 data, this study comprehensively analyzed the burden of kidney cancer attributable to high BMI among middle-aged males in East and Southeast Asia and 20 countries over 32 years, projecting trends to 2040. Results revealed accelerated disease burden growth exceeding global averages: by 2021, DALYs in East Asia approached 6.3 times the 1990 level, with an ASDR surge of 173.65%, while Southeast Asia saw DALYs rise to 5.8 times and ASDR increase by 123.75%. Concurrently, mortality and ASMR also rose significantly, underscoring the persistent challenge in controlling high BMI-related kidney cancer in these regions. This escalation may stem from improved early screening technologies and expanded coverage, enhancing detection of kidney cancer cases. Furthermore, rapid socioeconomic growth has amplified access to processed foods high in salt, fat, and sugar, while sedentary lifestyles have become a key driver of high BMI ^[25]^.

Our study reveals a higher burden of kidney cancer attributable to high BMI among middle-aged males in East Asia compared to Southeast Asia. In 2021, East Asia exhibited an ASDR of 15.63 (95% UI: 5.96–28.01) per 100,000 with an EAPC of 3.772 (95% UI: 3.538–4.006), while Southeast Asia showed an ASDR of 9.4 (95% UI: 3.52–15.45) per 100,000 and EAPC of 2.317 (95% UI: 2.115–2.519). A similar trend was observed for ASMR. This disparity may be attributed to East Asia’s higher number of developed regions, where research confirms a positive correlation between kidney cancer mortality and the Human Development Index (HDI) ^[26]^. Consistently, our findings demonstrate that ASDR and ASMR increase with higher SDI levels across both regions. Moreover, East Asian populations exhibit distinct physiological susceptibilities—including propensity for visceral adiposity, insulin resistance, and diabetes—that potentially amplify metabolic risks ^[27]^.

Notably, significant disparities in the disease burden of kidney cancer attributable to high BMI exist across East and Southeast Asia due to variations in climate, lifestyle, and economic systems. In 2021, East Asia exhibited the highest burden, with China accounting for the largest male population and the highest DALYs (27,276.85) and Deaths (704.26), alongside elevated ASDR (15.54 per 100,000) and ASMR (0.40 per 100,000), as well as a steep upward trend. This aligns with China’s high obesity prevalence (overweight: 34.3%; obesity: 16.4% during 2015–2019) ^[28]^. In contrast, Southeast Asia showed divergent patterns: Indonesia reported the highest regional DALYs (2,108.42) and Deaths (55.19) but lower ASDR (8.72 per 100,000) and ASMR (0.23 per 100,000). A notable anomaly occurred in Sri Lanka, where ASDR and ASMR plummeted sharply between 2002–2005, likely due to disrupted healthcare services amid prolonged conflicts in its northern and eastern regions. Collectively, these trends underscore how political instability, economic pressures, and social inequities persistently exacerbate disease burden disparities across these regions ^[29, 30]^.

Joinpoint analysis revealed that middle-aged males aged 55–59 years bear the highest burden of kidney cancer attributable to high BMI in East and Southeast Asia. This aligns with evidence from China, where 58.9% of new cases and 77.9% of deaths occur in individuals aged ≥55 years ^[31]^. However, in East Asia, the growth rates of ASDR and ASMR were lower in the 55–59 age group compared to the 45–49 cohort, likely due to advanced medical technologies and widespread health education facilitating early diagnosis and effective treatment. Furthermore, a decline in burden among East Asians aged 45–54 was observed during 2012–2015, coinciding with the implementation of China’s National Chronic Disease Prevention and Control Plan (2012–2015), which targeted obesity control and enhanced early screening capabilities in primary healthcare settings. In contrast, Southeast Asia exhibited sustained upward trends across all age groups, except for a transient decline in the 50– 54 cohort during 2001–2005. This anomaly likely stems from healthcare disruptions and data collection challenges during regional conflicts (e.g., political instability in Indonesia, the Philippines, and Thailand), natural disasters (e.g., the 2004 Indian Ocean tsunami that destroyed 80% of healthcare facilities in Aceh, Indonesia), and public health crises (e.g., the 2002 SARS outbreak that strained medical resources across Southeast Asia), collectively reflecting the region’s complex challenges in security, governance, and health system resilience.

Significant health inequities in East and Southeast Asia are exacerbated by socioeconomic disparities, cultural practices, and unequal healthcare resource distribution ^[32, 33]^. Projections indicate that by 2040, the burden of kidney cancer attributable to high BMI among middle-aged males in these regions will escalate, with ASDR and ASMR surpassing global averages—notably higher ASDR growth in East Asia and accelerated ASMR increases in Southeast Asia. To effectively mitigate this burden, a multidimensional strategy is imperative: (1) Enhance weight management through reduced abdominal adiposity via low-salt, low-fat diets and ≥150 minutes/week of moderate-intensity exercise to lower obesity-associated risks ^[34]^; (2) Scale early screening by integrating annual urinary ultrasonography and urinalysis for high-risk groups (e.g., elevated BMI/abdominal obesity), embedding BMI into routine assessments to improve early diagnosis ^[35]^; (3) Optimize healthcare resource allocation through regional cooperation to bridge diagnostic-treatment gaps (e.g., expanding radiotherapy access in Indonesia and Vietnam) and enhance basic care accessibility in low-SDI areas to reduce high mortality; (4) Target high-risk males aged 40–54 with tailored smoking cessation, blood pressure management, and obesity interventions to attenuate incidence acceleration ^[36]^; and (5) Standardize protocols and transnational resource-sharing by unifying diagnostic criteria and upskilling primary healthcare workers to balance screening coverage with therapeutic access, ensuring sustainable burden reduction.

This study has several limitations. Firstly, variations in diagnostic criteria across countries challenge precise cross-national comparisons of kidney cancer burden. Furthermore, the absence of incidence and prevalence data in the GBD database constrained comprehensive burden analysis. Additionally, our assumption of BMI as an independent risk factor may overlook potential interactions with other variables.

## Conclusion

The epidemiological burden of kidney cancer attributable to high BMI among middle-aged males in East and Southeast Asia demonstrates a significant upward trend across all age groups, with growth rates far exceeding global averages. Significant health inequalities exist between regions, with East Asia bearing a higher burden than Southeast Asia, and high SDI regions experiencing disproportionately greater impacts. Projections indicate a continued rise in this burden by 2040, underscoring the urgent need to develop and implement targeted interventions addressing the escalating obesity epidemic and its effect on kidney cancer. This is particularly critical for middle-aged males and high-burden regions to mitigate the growing kidney cancer burden.

## Abbreviations

GBD: global burden of disease
BMI: body mass index
DALYs: disability-adjusted life years
ASR: age-standardized rate
ASDR: age-standardized DALY rate
ASMR: age-standardized mortality rate
SDI: socio-demographic index
BAPC: Bayesian age-period-cohort
EAPC: estimated annual percentage change
AAPC: average annual percentage change
UI: uncertainty interval

## Acknowledgements

We sincerely acknowledge the contributions of database researchers whose meticulously developed resources have advanced progress in public health. Their sustained efforts to improve global health data represent an indispensable asset to the scientific community.

## Data availability

All data used in this study were obtained from the GBD 2021 database (https://vizhub.healthdata.org/gbd-results/).

## Ethics approval and consent to participate

This is a retrospective, observational cohort study utilizing publicly available database resources; therefore, the requirement for informed consent was waived. The consent was not applicable. All authors have read and agreed to the published version of the manuscript.

## Supporting information

**S1 Table. Detailed data of kidney cancer attributable to high BMI among middle-aged males in East and Southeast Asia from 1990 to 2021.**

**S2 Table. Detailed data from Joinpoint analysis.**

**S3 Table. Detailed data from Bayesian age-period-cohort analysis.**

## Author Contributions

W.S.: Conceptualization, Funding Acquisition, Project Administration, Resources, Supervision, Writing-Review & Editing. Z.W.: Conceptualization, Data Curation, Formal Analysis, Methodology, Software, Visualization, Writing-Original Draft Preparation.Y.W., Q.W., and Y.R.: Writing-Review & Editing.

